# Multi-task learning for activity detection in neovascular age-related macular degeneration

**DOI:** 10.1101/2022.06.13.22276315

**Authors:** Murat Seçkin Ayhan, Hanna Faber, Laura Kühlewein, Werner Inhoffen, Gulnar Aliyeva, Focke Ziemssen, Philipp Berens

## Abstract

**Purpose:** Comparison of performance and explainability of a multi-task convolutional deep neuronal network to single-task networks for activity detection in neovascular age-dependent macular degeneration.

**Methods:** From n = 70 patients (46 female, 24 male) who attended the University Eye Hospital Tübingen 3762 optical coherence tomography B-scans (right eye: 2011, left eye: 1751) were acquired with Heidelberg Spectralis, Heidelberg, Germany. B-scans were graded by a retina specialist and an ophthalmology resident, and then used to develop a multi-task deep learning model to predict disease activity in neovascular age-related macular degeneration along with the presence of sub- and intraretinal fluid. We used performance metrics for comparison to single-task networks and visualized the DNN-based decision with t-distributed stochastic neighbor embedding and clinically validated saliency mapping techniques.

**Results:** The multi-task model surpassed single-task networks in accuracy for activity detection (94.2). Furthermore, compared to single-task networks, visualizations via t-distributed stochastic neighbor embedding and saliency maps highlighted that multi-task networks’ decisions for activity detection in neovascular age-related macular degeneration were highly consistent with the presence of both sub- and intraretinal fluid.

**Conclusions:** Multi-task learning increases the performance of neuronal networks for predicting disease activity, while providing clinicians with an easily accessible decision control, which resembles human reasoning.

**Translational Relevance:** By improving nAMD activity detection performance and transparency of automated decisions, multi-task DNNs can support the translation of machine learning research into clinical decision support systems for nAMD activity detection.

## 1 Introduction

Neovascular age-related macular degeneration (nAMD) is a sight-threatening disease and a common cause of vision loss worldwide.^1–3^ Among the basic features of nAMD are subretinal fluid (SRF) and intraretinal fluid (IRF), which serve as surrogate markers of nAMD activity and can be monitored using optical coherence tomography (OCT)^4,5^ (Fig. 1).

**Figure 1:**
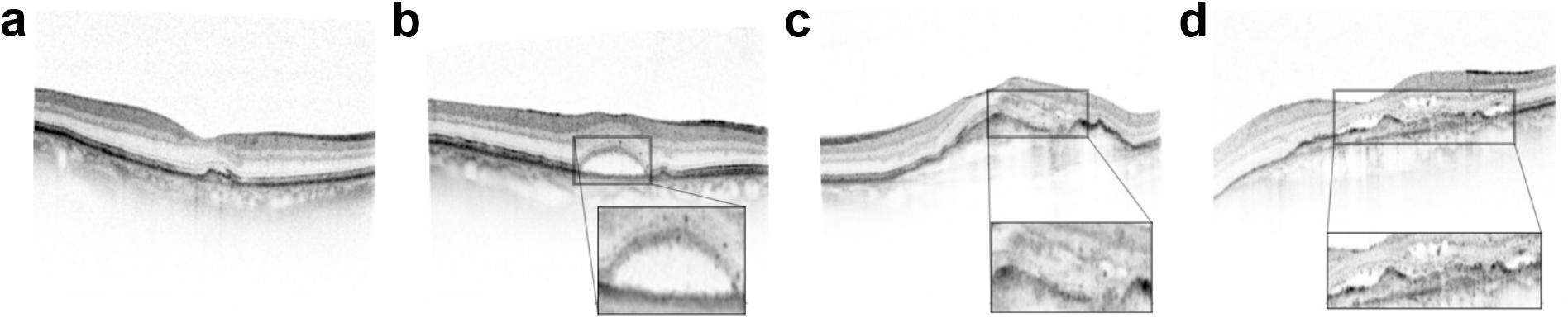
Exemplary retinal images (B-scans) with neovascular age-related macular degeneration (nAMD). Solid and dotted arrows indicate subretinal and intraretinal fluid, respectively. **(a)**: no nAMD activity. **(b)**: nAMD activity due to subretinal fluid (SRF). **(c)**: nAMD activity due to intraretinal fluid (IRF). **(d)**: nAMD activity due to both SRF and IRF.

In nAMD, increased levels of vascular endothelial growth factor (VEGF) lead to formation of new vessels from the choroidal and/or retinal vasculature. If leakage from these vessels exceeds local clearance rates, fluid builds up, leading to IRF and SRF.^4^ IRF is assumed to originate from vascular leakage from intraretinal neovasculaturisation and/or retinal vasculature or from diffusion through the outer retina due to changes within the external limiting membrane.^4^ In contrast, SRF formation likely results from malfunction of the retinal pigment epithelium with reduced removal rates.^4^ Due to the partially different pathophysiology, IRF and SRF can occur both simultaneously and independently from each other.^4,6^ In addition, the characterisation of the lesion based on IRF and SRF could help to determine visual outcome.^7^

Treatment with intravitreal anti-VEGF agents efficiently restores the balance between fluid formation and retinal removal and is standard of care, when IRF or SRF in nAMD is detected via OCT.^5^ Prompt treatment initiation is necessary to prevent vision loss.^8–10^ Additionally, this chronic disease demands high-frequent therapy monitoring, which has put considerable burden on patients, their families and ophthalmological care since its initial approval in 2006.^11–14^ Since the number of patients suffering from AMD is thought to rise from 196 million in 2020 to 288 million in 2040, the care needed will also rise.^2^ Hence, automated solutions making the diagnostic processes more efficient have considerable appeal. For example, deep neural networks (DNNs) have been used for automatic referral decisions^15^ and predicting disease conversion to nAMD.^16^ Automated algorithms could detect both SRF and IRF more reliably than retinal specialists especially in less conspicuous cases.^17^ Ideally, such automated tools serve to support retinal specialists in their decision making. In collaboration, a retina specialist assisted by an artificial intelligence (AI) tool can outperform the model alone, e.g., for the task of diabetic retinopathy grading.^18^ To this end, computational tools need to explain their decisions and communicate their uncertainty to the treating ophthalmologist.^19, 20^

Here, we develop a convolutional deep learning model based on the concept of multi-task learning.^21, 22^ Multi-task learning is a generalization of the widely used single-task learning, where models are trained for multiple input-output mappings simultaneously (Fig. 2). For instance, multi-task models can be used to capture different characteristics of dry AMD, such as drusen area, geographic atrophy, increased pig-ment, and depigmentation, to combine these outputs into final AMD diagnosis w.r.t. a 9-step severity scale.^23^ Multi-task learning has also shown prognostic value when applied to survival analysis via two simultaneous prediction tasks: drusen and pigmentation grading.^24^ In a similar vein, our multi-task model detects SRF, IRF and nAMD activity in parallel. However, it generates distinct outputs for each of these tasks and offers well-calibrated uncertainty estimates for each of them, which is unique to our study. As the fluid compartment plays a decisive role in the treatment outcome^25–27^ with the simultaneous presence of IRF and SRF being associated with the worst prognosis,^9^ we visualize the representation driving the DNN-based decisions using t-distributed stochastic neighbor embedding (t-SNE)^28,29^ and investigate the model’s decisions using clinically validated saliency mapping techniques.^30^ Thus, together with well-calibrated uncertainty reports, our work provides an interpretable tool for the ophthalmologist to rapidly access the neural network’s decision process on both population-based and individual-patient levels as a prerequisite for clinical application.

**Figure 2:**
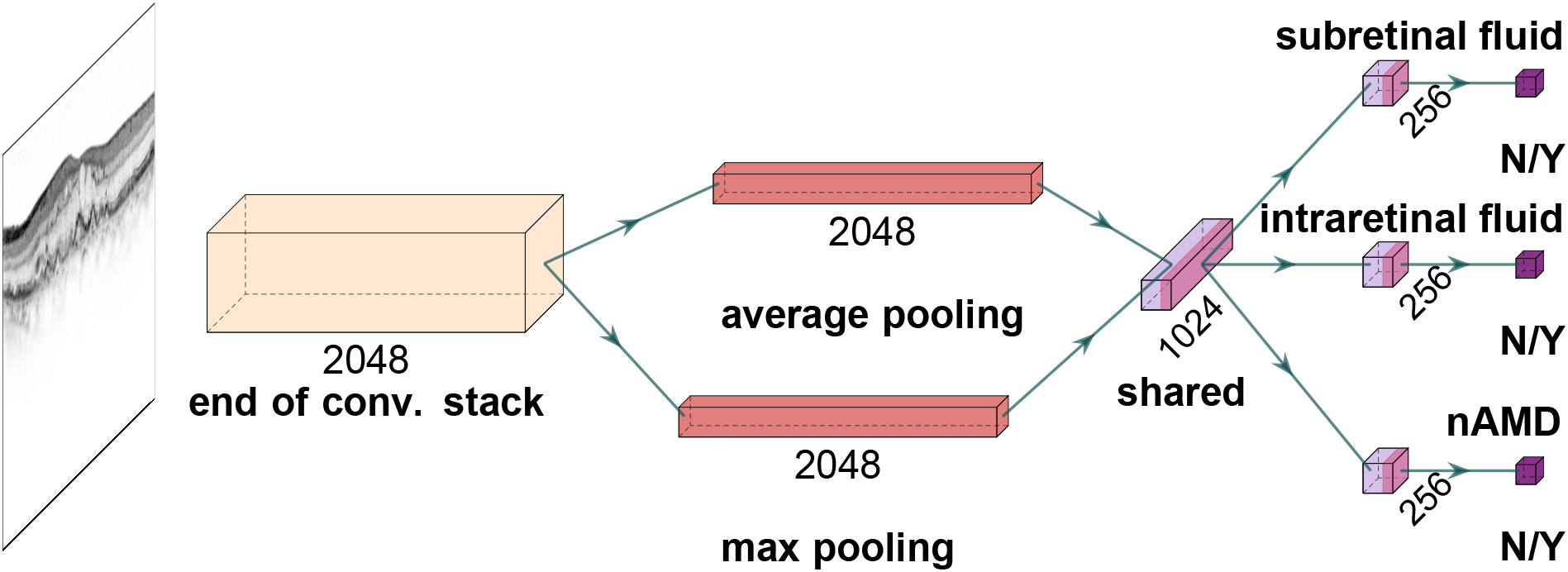
A deep neural network for simultaneous detection of subretinal and intraretinal fluid as well as the nAMD activity from OCT B-scans. Given a B-scan, convolutional stack of the InceptionV3 architecture extracts 2048 feature maps. These are average and max pooled, and fed into a fully connected (dense) layer with 1024 units for shared representation. Then, task-specific heads specialize into individual tasks and single units with sigmoid function achieve binary classification based on 256 task-specific features.

## 2 Methods

### 2.1 Data Collection

This study included 70 patients (46 females, 24 males) with nAMD at least in one eye, seen by an ophthalmologist resident (AG) in the Macula clinic at the University Eye Hospital Tübingen. Exclusion criteria were any other cause of neovascularisation, any coexisting retinal pathology (e.g. epiretinal membrane, macular hole, diabetic retinopathy), glaucoma and media opacity preventing sufficient image quality.

3762 B-scans (2011 right eye, 1751 left eye) of 440 × 512 pixels with Heidelberg Spectralis OCT (Heidelberg Engineering, Heidelberg, Germany) were included in the study. A retina specialist of the same hospital (IW) assessed disease activitiy and presence of IRF and SRF on each individual B-Scan. (Fig. 1). Disease activity was also graded by a resident (AG). B-scans were assigned to a training, validation or test set (Table 1). All images of one patient were assigned to one set to avoid information leakage. The study was conducted in accordance with the tenets of the Declaration of Helsinki and approved by the local institutional ethics committee of the University of Tübingen, which waived the requirement for patient consent due to the study’s retrospective character.

**Table 1:** OCT Data distribution of subretinal fluid (SRF), intraretinal fluid (IRF) and active nAMD in B-Scans in training, validation and test sets, respectively. Absolute and relative numbers are shown.

|  | Training |  |  | Validation |  |  | Test |  |  |
| --- | --- | --- | --- | --- | --- | --- | --- | --- | --- |
|  | *Subretinal fluid* | *Intraretinal fluid* | *Active nAMD* | *Subretinal fluid* | *Intraretinal fluid* | *Active nAMD* | *Subretinal fluid* | *Intraretinal fluid* | *Active nAMD* |
| Yes | 639<br>(0.232) | 286<br>(0.104) | 848<br>(0.308) | 69<br>(0.170) | 58<br>(0.143) | 101<br>(0.248) | 161<br>(0.267) | 153<br>(0.253) | 269<br>(0.445) |
| No | 2112<br>(0.768) | 2465<br>(0.896) | 1903<br>(0.692) | 338<br>(0.830) | 349<br>(0.857) | 306<br>(0.752) | 443<br>(0.733) | 451<br>(0.747) | 335<br>(0.555) |

### 2.2 Diagnostic Tasks, Network Architecture and Model Development

We developed a multi-task DNN to detect the presence of SRF and IRF as well as the nAMD activity from OCT B-scans (Fig. 2). As backbone, we used the InceptionV3 architecture^31^ via Keras,^32^ which was pre-trained on ImageNet^33^ for 1000-way classification via a softmax function. We used the InceptionV3 DNN’s convolutional stack as is but linked max pooling and average pooling layers to the end of convolutional stack and concatenated their outputs to obtain 4096-dimensional feature vectors. These were followed by a dense layer, which yielded a shared representation with 1024 features. To this, we added task-specific heads with 256 units, which specialized into their respective tasks. Then, task-specific binary decisions were achieved by single units equipped with sigmoid functions. For training our DNNs in both single and multi-task scenarios, we resorted to the retina specialist’s set of labels.

We trained our networks with equally weighted cross-entropy losses for all tasks on the training images: 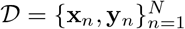, where **y**_*n*_ was a vector of binary labels indicating nAMD activity and the presence of IRF or SRF in an image **x**_*n*_. Parameterized by *θ*, a DNN *f*_*θ*_(·) was optimized with respect to the total cross-entropy on the training data:

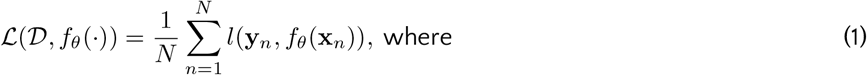

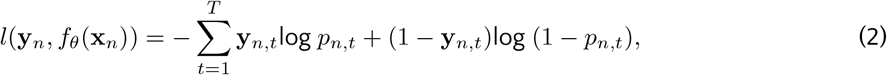

*p*_*n,t*_ was a probability estimated via the sigmoid function for a task indicated by *t*, and *T* was the total number of tasks. For *T* = 1, multi-task learning was reduced to single-task learning based on the same architecture but with only one task head. We also developed a 2-task model to perform the SRF and IRF detection tasks (*T* = 2), while eliminating the redundancy of the nAMD activity detection task, which is, in principle, a function of the former two.

To address the class imbalance (Table 1), we used random oversampling (see Section 2.2.2 for details). We trained the DNN using Stochastic Gradient Descent (SGD) with Nesterov’s Accelerated Gradients (NAG),^34,35^ minibatch size of eight, a momentum coefficient of 0.9, an initial learning rate of 5·10^*−*4^, a decay rate of 10^*−*6^ and a regularization constant of 10^*−*5^ for 120 or 150 epochs (see Section 2.2.1 for longer training). During the first five epochs, the convolutional stack was frozen and only dense layers were trained. Then, all layers were fine-tuned to all tasks. The best models were selected based on total validation loss after each epoch and used for inference on the test set.

#### 2.2.1 Data augmentation and preprocessing

We used *mixup*^36^ for data augmentation during training. Mixup generates artificial examples through the convex combinations of randomly sampled data points. We adapted *mixup* to our multi-task learning scenario as follows:

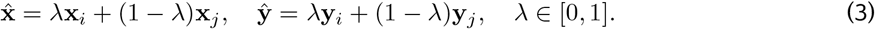

Mixing was controlled by *λ* ∼*Beta*(*α, α*), where *α* ∈ (0, ∞). For *α* = 0, *λ* is either 0 or 1, and there is no mixing. We used 0, 0.05 0.1, and 0.2 for *α* and trained networks for 120 epochs when not mixing and 150 epochs when mixing. Also, to allow for a warm-up period when mixing,^36^ we set *α* = 0 for the first five epochs.

In addition, we applied common data augmentation operations such as adjustment of brightness within ±10%, horizontal and vertical flipping, up and down scaling within ±10%, translation of pixels horizontally and vertically within ±30 positions and random rotation within ±45 degrees. After all data augmentation operations, we used an appropriate preprocessing function^1^ from the Keras API.^32^

#### 2.2.2 Quantification of uncertainty via *mixup* and Deep Ensembles

DNNs often do not generate well-calibrated and reliable uncertainty estimates for their decision.^37–41^ How-ever, quantification of diagnostic uncertainty is crucial for treatment decisions since proper management can minimize diagnostic errors, delays or excess healthcare utilization.^42^ *mixup*^36^ improves the calibration of DNN outputs by smoothing labels through their convex combinations (Eq. 3).^43^ In addition, we used Deep Ensembles^39^ consisting of multiple DNNs with different random initializations.^39,44^ This can improve upon the single network performance both in accuracy and calibration, even with small numbers of DNNs.^39,44–46^ We used ensembles with three DNNs, for which we enforced diversity by a specialized oversampling strategy: for each DNN, we oversampled training images with respect to one of the task’s labels. This enabled DNNs to train on a balanced dataset while also learning about other tasks, even though the data were not balanced for these. We then used the ensemble’s mean output for predictions and quantified uncertainty in terms of entropy, given the average predictive probabilities.

### 2.3 Low-dimensional embedding of images

We used t-SNE^28^ to obtain further insights into the decision-making process of our ensemble model. t-SNE is a non-linear dimensionality reduction method, that embeds high-dimensional data points into a low-dimensional space. We concatenated features from ensemble members’ predetermined read-out layers and performed t-SNE based on them, embedding each B-scan into the two-dimensional plane. We used *openTSNE*^47^ with PCA initialization to better preserve the global structure of the data and improve the reproducibility.^29^ A perplexity of 200 for 1500 iterations with an early exaggeration coefficient of 12 for the first 500 iterations was used according to best-practice strategies.^29^ Similarities between data points were measured by Euclidean distance in the feature space.

### 2.4 Saliency Maps

We used Layer-wise Relevance Propagation (LRP)^48^ to compute saliency maps highlighting the regions in the OCT images which contributed to the DNN decisions, as it provides most clinically relevant tasks.^30^ We created three saliency maps for each OCT slice: subretinal (cyan), intraretinal (magenta) and diesease activity in nAMD (yellow) (Fig. 5). To improve the visualization of the salient regions, saliency maps were postprocessed.^30^ Saliency maps were only shown for predictions with an estimated probability greater than 0.5 since previous work has shown, that especially in absence of disease, saliency maps can lead physicans to overdiagnosis.^18^

## 3 Results

We developed an ensemble of three multi-task DNNs to simultaneously detect SRF, IRF and activity of nAMD on OCT B-scans (Fig. 1). Each DNN consisted of a shared convolutional core combined with pooling operations and a fully connected (dense) layer (Fig. 2). The resulting shared representation served as the basis for the decisions of the three task-specific heads. The idea behind this approach is that the DNN can benefit from the shared representation induced by combining information from different tasks. We compared the performance of the multi-task model with more specialized single-task models, where we constructed three DNNs for each task, which did not share any representation but were trained independently. In addition, we also used a 2-task model that simultaneously detected only SRF and IRF, without the nAMD activity detection head.

All DNNs were trained on the same dataset (see Table 1 and Methods), which was graded according to the nAMD activity by a retina specialist (IW) and and an ophthalmologist resident (AG) with high inter-grader agreement on disease activity (Cohen’s kappa = 0.86). In a second step, the retina specialist further examined the data for the presence of IRF and SRF. The two retinal fluid types occurred largely independently, while there was natural overlap of both with the active AMD label (Table 2).

**Table 2:** Agreement of task-specific labels across training, validation and test sets, measured via Cohen’s kappa statistic, which is essentially a number between -1 and 1. While 1 indicates a full agreement, lower scores mean less agreement. Negative scores indicate disagreement.

|  | Training |  |  | Validation |  |  | Test |  |  |
| --- | --- | --- | --- | --- | --- | --- | --- | --- | --- |
|  | *Subretinal fluid* | *Intraretinal fluid* | *Active nAMD* | *Subretinal fluid* | *Intraretinal fluid* | *Active nAMD* | *Subretinal fluid* | *Intraretinal fluid* | *Active nAMD* |
| *Subretinal fluid* | - | -0.02 | 0.79 | - | 0.26 | 0.75 | - | -0.02 | 0.59 |
| *Intraretinal fluid* | -0.02 | - | 0.37 | 0.26 | - | 0.65 | -0.02 | - | 0.57 |

We selected the 3-task model with the best accuracy for the activity detection task on the validation set and report accuracy values computed on an independent test set (Table 3). The 3-task model was well calibrated on the test set (Adaptive expected calibration error^41^ of 0.0147 for SRF, 0.0104 for IRF and 0.0263 for active nAMD).

**Table 3:** Accuracy of ensembles for various degrees of mixing (indicated by *α*). Gray row indicates the ensemble of choice for further analysis based on the validation performance for the activity detection task. In the 2-task scenario, the average validation accuracy of SRF and IRF detection tasks was used for model selection.

| *Single task* |  |  |  |  |  |  |  |  |  |
| --- | --- | --- | --- | --- | --- | --- | --- | --- | --- |
|  | Training |  |  | Validation |  |  | Test |  |  |
|  | *Subretinal fluid* | *Intraretinal fluid* | *Active nAMD* | *Subretinal fluid* | *Intraretinal fluid* | *Active nAMD* | *Subretinal fluid* | *Intraretinal fluid* | *Active nAMD* |
| $\alpha = 0$ | 1.000 | 1.000 | 1.000 | 0.988 | 0.971 | 0.958 | 0.924 | 0.950 | 0.914 |
| $\alpha = 0.05$ | 0.983 | 0.994 | 0.975 | 0.971 | 0.963 | 0.951 | 0.906 | 0.919 | 0.909 |
| $\alpha = 0.1$ | 0.978 | 0.994 | 0.948 | 0.948 | 0.919 | 0.929 | 0.868 | 0.891 | 0.856 |
| $\alpha = 0.2$ | 0.983 | 0.991 | 0.851 | 0.975 | 0.946 | 0.853 | 0.881 | 0.909 | 0.702 |
| *Multiple tasks* |  |  |  |  |  |  |  |  |  |
| *SRF and IRF* |  |  |  |  |  |  |  |  |  |
| $\alpha = 0$ | 0.999 | 1.000 | - | 0.968 | 0.961 | - | 0.902 | 0.937 | - |
| $\alpha = 0.05$ | 1.000 | 0.999 | - | 0.983 | 0.966 | - | 0.927 | 0.919 | - |
| $\alpha = 0.1$ | 0.999 | 0.999 | - | 0.983 | 0.973 | - | 0.911 | 0.924 | - |
| $\alpha = 0.2$ | 0.999 | 1.000 | - | 0.983 | 0.963 | - | 0.917 | 0.932 | - |
| *SRF, IRF and nAMD activity* |  |  |  |  |  |  |  |  |  |
| $\alpha = 0$ | 1.000 | 0.995 | 0.998 | 0.973 | 0.973 | 0.961 | 0.914 | 0.935 | 0.940 |
| $\alpha = 0.05$ | 0.999 | 0.998 | 1.000 | 0.971 | 0.971 | 0.966 | 0.917 | 0.937 | 0.942 |
| $\alpha = 0.1$ | 1.000 | 0.997 | 0.998 | 0.983 | 0.968 | 0.966 | 0.916 | 0.957 | 0.939 |
| $\alpha = 0.2$ | 1.000 | 0.998 | 1.000 | 0.971 | 0.966 | 0.966 | 0.894 | 0.937 | 0.906 |

We found that the performance of the 3-task model surpassed the single-task model performance in disease activity detection, reaching an accuracy of 94.2 % for the multi-task model vs. 91.4% for the single task model (Table 3, Fig. 3). This 3-task model optimized for AMD activity detection performed slightly worse than the single-task models for SRF and IRF detection (SRF: accuracy of 0.917 vs. 0.924 for multi-task vs. single-task; IRF: 0.937 vs. 0.950). For the 2-task scenario, we selected the model with the highest average validation accuracy across the SRF and IRF detection tasks. Interestingly, the 2-task model performed worse than the single-task and 3-task models. This highlights the importance of the explicit nAMD activity detection head in the 3-task model.

**Figure 3:**
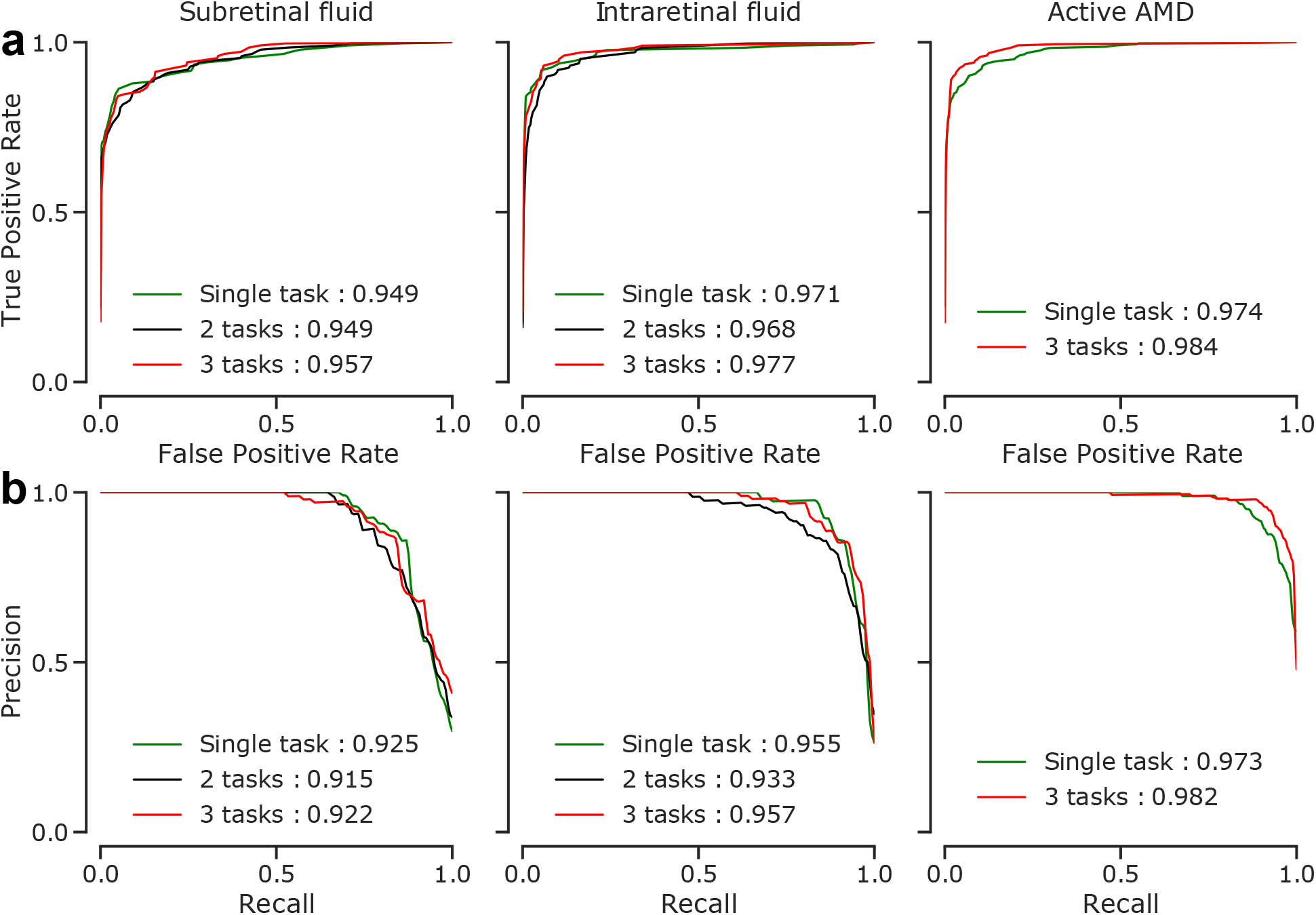
Performance curves of the selected models on the test images. Area under the curve (AUC) values given for models also summarize the overall performance into one number (higher is better). **(a)** Receiver Operating Characteristics (ROC) curves. **(b)** Precision-recall curves.

We then further studied the representations learned by the models to gain insight into their decision making-process. To this end, we extracted the representations of individual OCT scans from both single-task and multi-task models and created two-dimensional embeddings of these via t-SNE (Fig. 4. In these visualizations, each point represents an individual OCT scan. Scans which are similar to each other according to the learned representation are mapped to nearby points. Of note, distances and in particular the size of white space between clusters in t-SNE plots should be carefully interpreted.^29,49^

**Figure 4:**
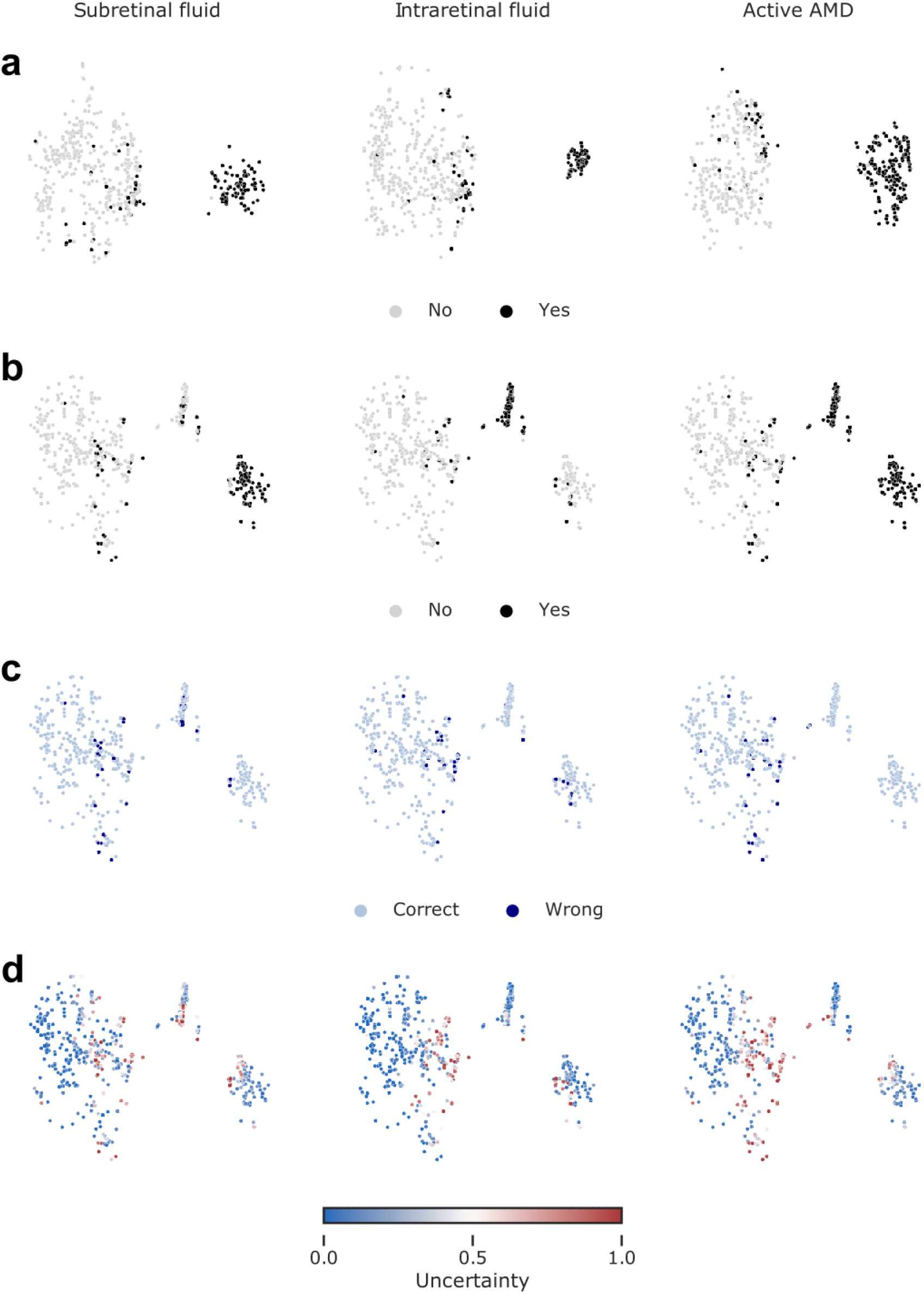
Visualization of data via t-SNE of ensemble-based representations. Only the test data are shown. **(a)** Low dimensional embedding of images based on the 1024-dimensional features from the prepenultimate layers of single-task networks. Colored with respect to the task-specific labels. **(b)** Same as in (a) but w.r.t. 1024 features from the shared representation layer of multi-task networks. **(c)** Same map as in (b) but colored w.r.t. correct and wrong predictions. **(d)** Same map as in (b) but colored w.r.t. uncertainty min-max normalized to [0, 1].

We labeled individual points according the evidence for SRF or IRF and overall AMD activity. In the single-taks DNNs, well-separated clusters were found, indicating only the learned task-label (Fig. 4a). For example, OCT scans with SRF present formed a single cluster, clearly distinct from the OCT scans without this label. In contrast, in the multi-task network subclusters within the active nAMD data points were observed (Fig. 4a, b): OCT scans labeled with SRF formed a well-separated cluster at the bottom right, as did scans with IRF labels at the top right (Fig. 4b). Interestingly, there was a small cluster in between these two which contained scans labeled with both. This suggests that multi-task DNNs learned a representation which could differentiate between the two fluid types. The few incorrectly classified OCT scans could be found within their clusters to be placed close towards other clusters (Fig. 4c) in areas where we also found examples with high classifier uncertainty (Fig. 4d).

We next studied how the multi-task representations emerged through processing in the network (Appendix, Fig. 7). While in the initial layers data points representing active nAMD were still uniformly distributed (Fig. 7, a-c), a clear separation of active nAMD cases developed gradually in later layers of the DNN (Fig. 7, d-g), leading to best separation in the shared representation (Fig. 7, h). The decision head for active AMD refined this representation only very little (Fig. 7, i).

We finally analyzed the saliency maps of the multi-task DNNs and asked whether the saliency maps for the subtasks of SRF and IRF detection obtained from the multi-task model allowed reasoning about evidence specific to these tasks. We generated saliency maps on four exemplary OCT scans using LRP^48^ (Figure 5). For an OCT scan with clearly active AMD and both SRF and IRF present (Figure 5a), we found that the active AMD saliency map focused on intraretinal fluids, which were also clearly visible in the task-specific saliency map, and faintly highlighted regions with SRF. The SRF saliency map, however, clearly highlighted SRF. In two further example scans with either IRF or SRF, respectively, active AMD saliency maps clearly corresponded to the individual task maps (Figure 5b,c). We also identified a rare failure case of the obtained saliency maps (Fig. 5d), where an OCT scan was falsely classified positive for SRF with a confidence of 0.614 due to the misclassification of IRF to SRF. We hypothesize that the DNN misclassified the superior border of the IRF as photoreceptor layer detached from the retinal pigment epithelium. The assumption that the DNN primarily recognizes contrast-rich interfaces such as SRF and IRF is further supported by the false labeling of cystoid spaces within choroid in Fig. 5b and d, while in a smoother, lower-contrast choroid saliency maps do not highlight any structures (Fig. 5. Comparision with salinecy maps from the single-task DNNs (Fig.6) to those generated from the multi-task models shows that those single-task saliency maps appear slightly more defined, but generally highlight similar areas.

**Figure 5:**
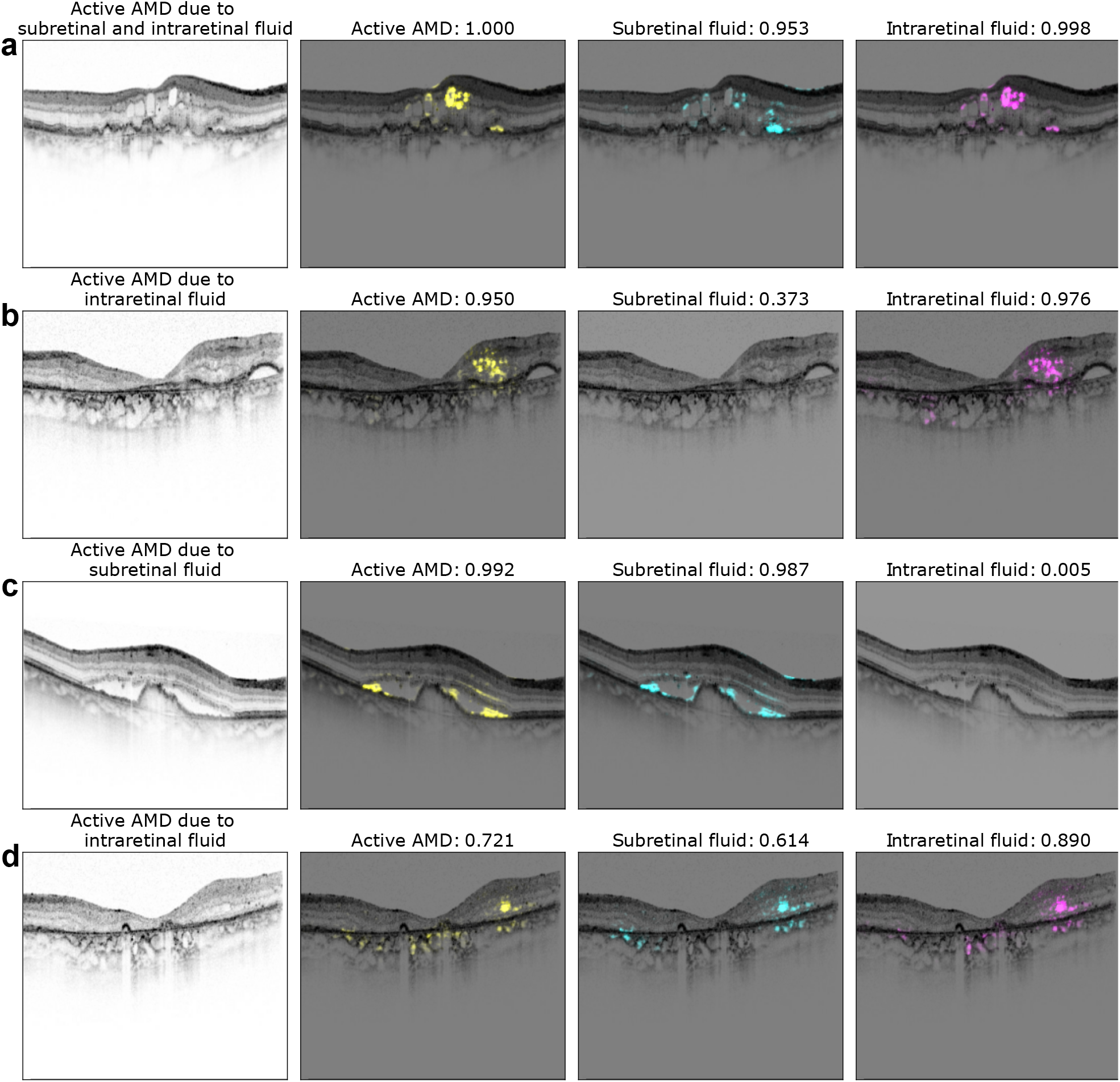
Exemplary saliency maps for four optical coherence tomography (OCT) images. The first column displays the OCT B-scan with the corresponding labeling of a retinal specialist. Second to fourth column show saliency maps and the network’s confidence for active nAMD (yellow), subretinal fluid (SRF) (cyan) and intraretinal fluid (IRF) (magenta). Note, that saliency maps are only shown in case of confidence *>* 0.5.

**Figure 6:**
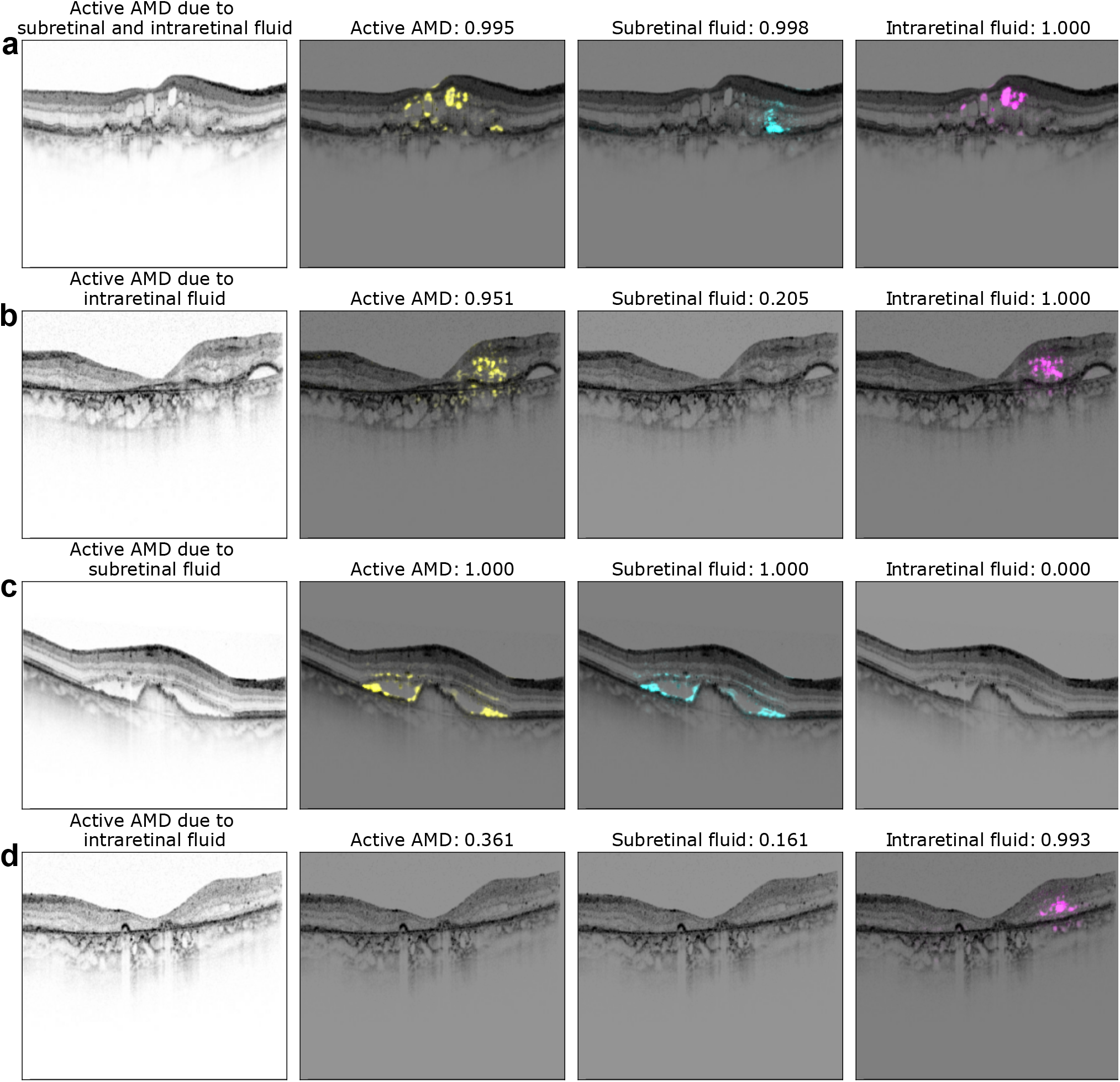
Exemplary saliency maps as in Fig. 5 but results were obtained from single-task models.

## 4 Discussion

In this study, we developed a multi-task learning model to simultaneously detect SRF, IRF as well as disease activity in OCT B-scans of nAMD patients. We showed that a 3-task model, which takes the presence of IRF and SRF into account to detect disease activity in nAMD, surpassed a single task model regarding accuracy in the activity detection task. Furthermore, our visualization of the multi-task model’s decision-making process via t-SNE showed that inactive and active nAMD B-scans formed different clusters. Among active AMD B-scans, three distinct clusters were observed, which contained OCT B-scans with either SRF, IRF or both fluid types. This separation could not be seen in the single-task models. Saliency maps of exemplary B-scans further corroborate that task-relevant information can be extracted from the multi-task networks.

Thus, multi-task DNN could serve as a basis for an explainable clinical decision support system for nAMD activity, providing support for clinicians in detecting active AMD, but would also allow clinicians to identify evidence in the relevant sub-tasks of finding SRF and IRF. A recent meta-analysis has provided evidence of varying influences of SRF and IRF on the visual outcome in nAMD patients.^50^ Stable SRF might not affect visual outcome, while fluctuations in IRF during treatment seem to negatively influence visual acuity.^50^ For this reason, treatment decisions in nAMD solely on a yes/no basis may not meet future treatment guidelines, which might rather require a sophisticated decision depending on the present fluid type and its variation in volume for or against an anti-VEGF injection.

Ophthalmology has recently seen a development of various artificial intelligence systems, yet their use in clinical routine remains rare with only few systems available on the market.^51, 52^ One big barrier is potential harm of the patient-physician relationship going hand in hand with the lack of trust in those systems.^53^ Here, we combined multi-task DNNs with different visualization methods to give an insight into the DNNs’ reasoning and increase transparency. First, we used t-SNE as visualization method for high-dimensional data^28,29^ (Fig. 4) to present the decision-making process of the model. This form of visualization provides an intuitively interpretable rationale for how OCT B-scans were graded by visualizing which other B-scans are similar. The results visualization may also increase an ophthalmologist’s confidence in the model since it illustrates shows that model’s decision making reasoning resembles their own (Fig. 7). In the future, the multi-task system could be extended for other signs of active nAMD such as hard exsudates, pigment epithelial detachment, subretinal hyperreflective materia or hyperreflective foci.^4^

We further analyzed the multi-task model’s decision on saliency maps of individual OCT-scans. Saliency maps highlight critical regions for the model’s decision and thus allow a quick visual control of its reasoning. This may be important in cases of advanced AMD, where fluid is due to degeneration rather than exudation to avoid overtreatment. However, different methods of saliency map agree to differing degrees with clinical annotations^30,54,55^ and saliency maps can lead to overdiagnosis.^18^ Therefore we used the saliency map technique with the best clinical relevance for AMD activity^30^ and displayed saliency maps in case of a confidence of the algorithm > 0.5. Compared to saliency maps of single task DNNs, multi-task saliency maps seem to draw slightly less sharp contours, however, we found good overlap between regions used for active AMD detection and those for SRF and IRF.

Future studies will need to assess how well these multi-task learning results transfer from this data sample acquired at a tertiary center in Germany. It would be desirable to perform similar analysis with larger and more diverse data sets, to thest also the generalization to other populations, different recording qualities as well as OCT devices (including mobile devices). Further, performance could be potentially increased by combining the multi-task network with a a segmentation layer,^15^ which could reduce false positive cases. Additionally, in clinical routine, activity decision is made on a whole volume not a single B-scan, which could technically be implemented by combining the results from individual B-scans, e.g. by majority voting or uncertainty propagation.

While the approval of anti-VEGF has decreased economic and overall treatment burden of nAMD measured in disability-adjusted life,^56,57^ a large number of patients still discontinues treatment.^58^ Patients named the need for assistance, either in the form of a travel companion or a family member, as the main reason for discontinuation.^14^ Additionally, recurrence of quiescent disease requiring prompt treatment is common, making life-long monitoring necessary.^59^ For these reasons, automated solutions allowing monitoring close home or even at home are promising technologies:^60,61^ They provide easier access and reduce the disease burden on the individual.^62^ Automated solutions for fluid detection have further gained popularity during the Covid-19 pandemic, which showed the devastating effects of delay or interruption of nAMD treatment on visual function.^9,59^ Despite promising results in laboratory settings, real-world data revealed significantly lower performance rates of home-based OCT with in particular SRF being overlooked by the system.^63^ This shows the necessity of further developments on the machine learning side to guarantee reliable use, with multi-task learning as suggested in this study being a viable option.

## Data Availability

The optical coherence tomography scans were obtained from the University Eye Clinic and their use was permitted by the Institutional Ethics Committee of the University of Tuebingen.

## 5 Acknowledgments

We thank the German Ministry of Science and Education (BMBF) for funding through the Tübingen AI Center (FKZ 01IS18039A) and the German Science Foundation for funding through a Heisenberg Professorship (BE5601/4-2) and the Excellence Cluster “Machine Learning — New Perspectives for Science” (EXC 2064, project number 390727645). H. Faber thanks the Faculty of Medicine, Eberhard Karls University of Tuebingen, Germany (application number 463–0–0) for additionally funding her research through the Junior Clinician Scientist Program (application number 463–0–0). We further thank Novartis AG for funding part of the research. The funding bodies did not have any influence in the study planning and design.

## Author contribution statement

MSA, HF and PB designed the research. MSA performed the experiments. GA, WI, FZ, LK were involved in data acuqisition. FZ, HF, GA, LK and WI provided medical advice. MSA, HF and PB wrote the manuscript with input from all authors. All authors approved the final version of the manuscript and agreed on being accountable for the work.

## Appendix

**Figure 7:**
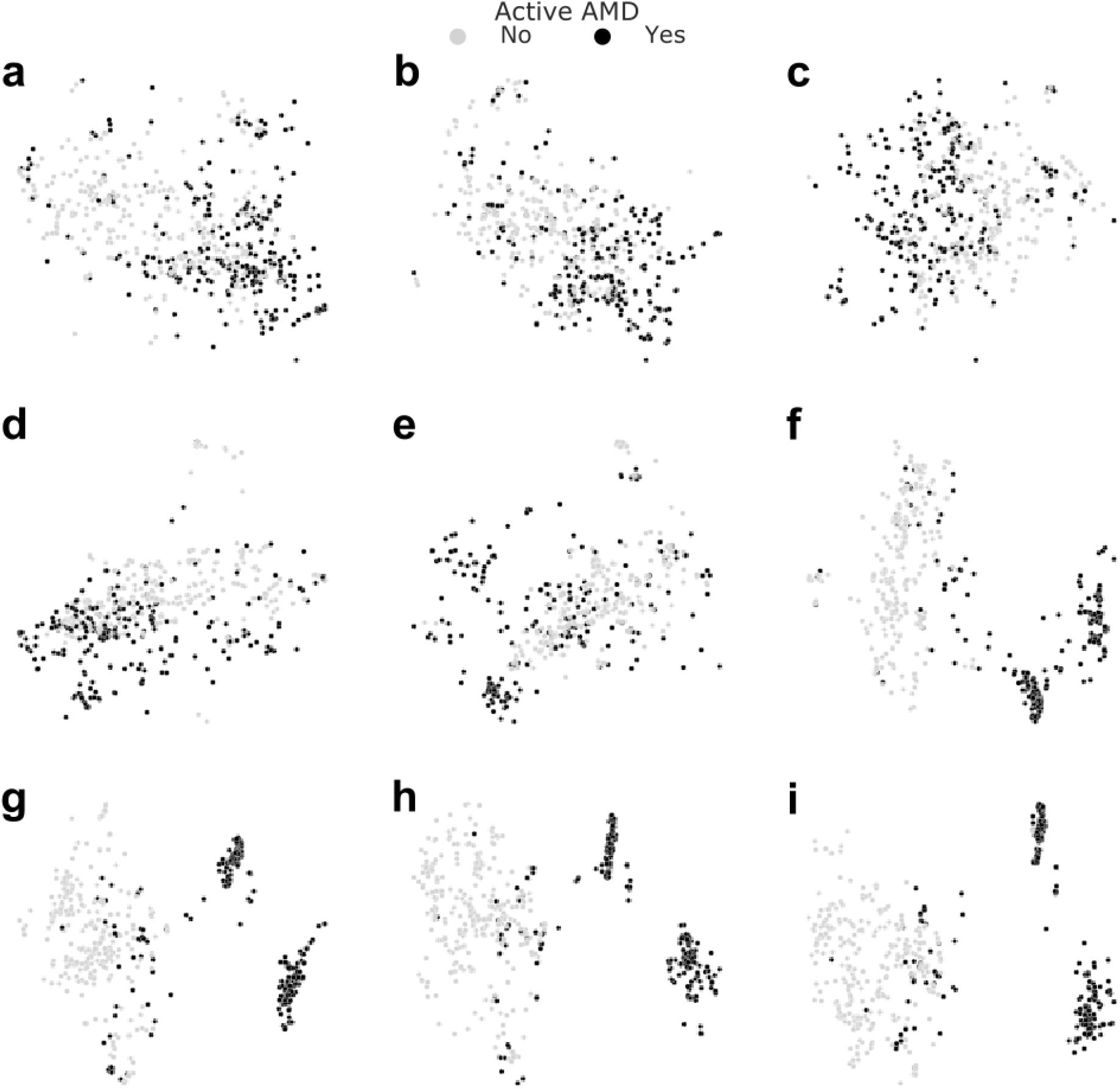
Layer-wise visualization of test data via t-SNE. Starting just before the first Inception module **(a)** and reading out feature representations yielded by every other module **(b-f)** along with the last Inception module **(g)**, the shared representation layer **(h)** and the nAMD activity detection head’s penultimate layer **(i)**, we performed t-SNE with the aforementioned settings. Useful representations emerged towards the end of convolutional stack and the task-specific representation allowed the best separation of nAMD active cases from those inactive. Exact read-out locations can be found in Fig. 8.

**Figure 8:**
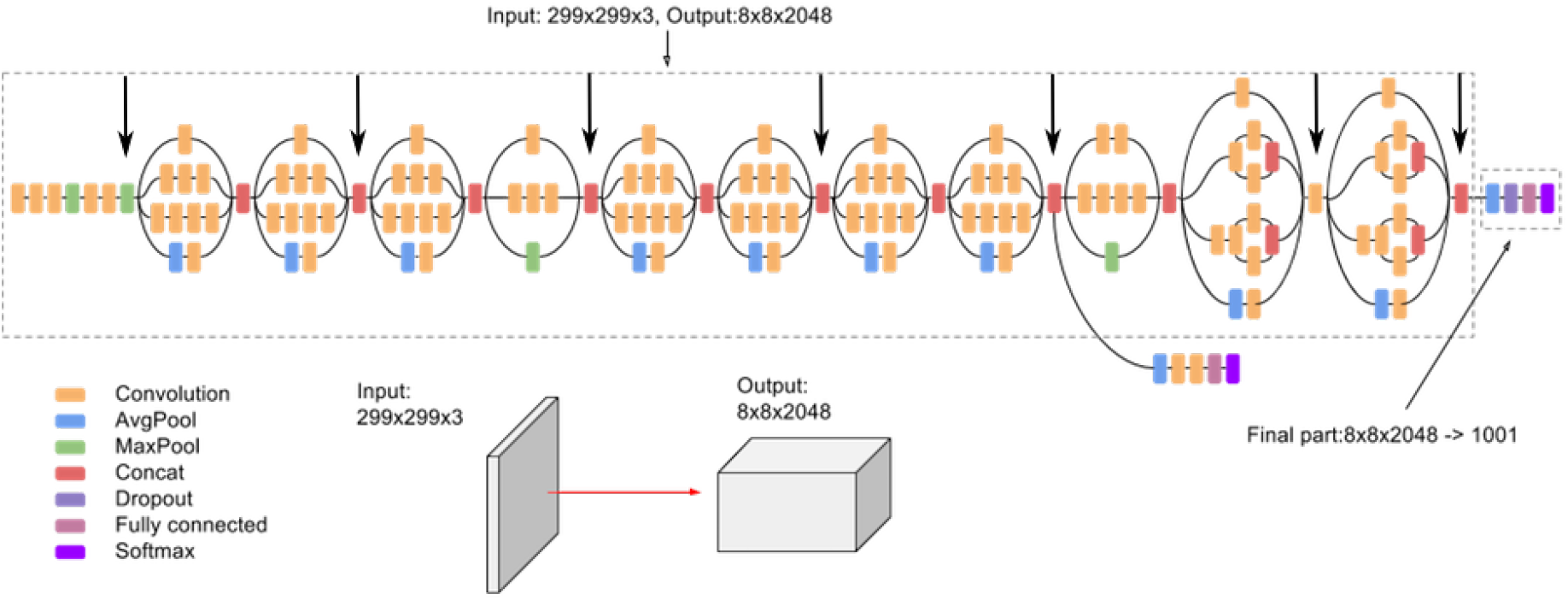
Read-out locations within the convolutional stack of the InceptionV3 architecture (indicated by big black arrows). In addition to these, we used the shared representation layer and task-specific layers of our multi-task networks (see Fig. 2). Base figure was obtained from https://cloud.google.com/tpu/docs/inception-v3-advanced.

## Footnotes

1 `keras.applications.inception_v3.preprocess_input`

